# EARLY SURGICAL OUTCOMES OF CHILDREN WITH WILMS’ TUMOR AT A TEACHING HOSPITAL IN WESTERN KENYA

**DOI:** 10.1101/2020.07.03.20145920

**Authors:** Nancy Kerubo Nyakundi, Pius Musau, Peter Saula

## Abstract

**Purpose:** To evaluate the early surgical outcomes of children with Wilms’ Tumour at a teaching hospital in Western Kenya.

**Patients and Methods:** A cross-sectional study was conducted among children diagnosed with Wilms’ tumour at Moi Teaching and Referral Hospital over a one-year period. The participants’ data on sociodemographic and clinical characteristics were collected using an interviewer administered questionnaire. Data on preoperative chemotherapy cycles, staging and predominant histological component, tumour rupture, early postoperative complications, duration of stay in surgical unit and mortality were obtained through chart reviews. Statistical association between patient characteristics and surgical outcomes were done.

**Results:** Thirty children with Wilms’ tumour were recruited into the study, with a mean age of 3.8 (SD± 1.5) years and a female to male ratio of 1.7:1. They all presented with an abdominal mass. The median duration of symptoms was 8 weeks (IQR 4, 12) prior to admission. Over half (53.3%) of the participants received 6 cycles of neoadjuvant chemotherapy while the rest received more. Two thirds of the tumours were on the left side while 53% of all tumours were classified as intermediate risk tumours. The early postoperative complications were intestinal obstruction (6.7%), surgical site infection (3.3%) and tumour rupture (3.3%). Post-operatively, patients stayed an average of 6.5 (SD±1.6) days in the surgical ward. There was a statistically significant association between duration of symptoms and surgical ward stay period (p=.044). There was no correlation between sociodemographic characteristics, clinical staging, and post-operative complications. None of the children died peri-operatively.

**Conclusions:** The Early surgical outcomes were favourable despite late presentation that increased the duration of stay in the surgical ward.

## Background

Wilms’ tumour (nephroblastoma) is the most common childhood kidney tumour as it accounts for approximately 91% of all kidney tumours^1^.It represents the most common abdominal tumour in infants and children in sub-Saharan Africa^2^. The treatment of nephroblastoma has been considered one of the greatest success stories in modern medicine. Its management in many low and middle-income countries (LMICs) consists of neo-adjuvant chemotherapy followed by radical nephroureterectomy, adjuvant chemotherapy and in some cases radiotherapy.

Despite advances in adjuvant therapy^3^, surgery is the mainstay oncologic treatment and is also critical for proper staging of the disease which aids in directing adjuvant therapy^4^. However, surgical complications are a recognized morbidity of the treatment of patients with Wilms’ tumour^5^ and their management remains an important part of paediatric oncologic treatment^3^. Wilms’ tumour is a common malignancy among the pediatric population under the age of 7 in our setting^1, 6^. This study evaluated the early surgical outcomes in patients who had undergone radical nephroureterectomy for Wilms’ Tumor at MTRH and the factors associated with the surgical outcomes.

## Patients and Methods

This was a descriptive cross-sectional study conducted at the Moi Teaching and Referral Hospital (MTRH) paediatric oncology and paediatric surgical wards. The hospital has a multidisciplinary medical team for the management of Wilms’ tumours. The participants were patients who presented with Wilms’ tumour and underwent radical nephrouretectomy at MTRH during the study period between July 2017 and June 2018. The study received prior ethical approval from the MU/MTRH Institutional Research and Ethics Committee (IREC) and parental consent was obtained for all the children enrolled.

The principal investigator reviewed each patient’s file a day prior to scheduled surgery to gather clinical characteristics such as age, duration of symptoms, preoperative chemotherapy cycles and review of imaging. Intraoperative findings were collected during surgery while postoperative complications were noted over the duration of stay in the surgical ward. Histopathology results gave the pathological staging and histological subtypes. Associations between the patient variables and the outcomes were done using Fishers’ Exact Test and Spearman’s Correlation Test.

## Results

A total of 30 children with Wilms’ tumour were recruited into the study. The participants’ age ranged from 2 to 7 years with a mean age of 3.8 years (±1.5) and a female to male ratio was 1.7:1. All the participants presented with abdominal mass, 14 (46.7%) had fever while 2 (6.7%) presented with macroscopic hematuria. The median duration of symptoms was 8 weeks (IQR 4, 12). Over half (53.3%) of the participants received 6 cycles of chemotherapy prior to surgery with the rest receiving seven (30%) and eight (16.7%) cycles. Two thirds of the tumours were on the left side while more than half (53%) of all tumours were classified as intermediate risk tumours. Early postoperative complications were intestinal obstruction (6.7%), surgical site infection (3.3%) and tumour rupture (3.3%) The mean duration of stay in the surgical ward was 6.5 (SD±1.6) days (Table 1). There was a statistically significant association between duration of symptoms and stay in the surgical ward (p=0.044) as shown in Table 2. Furthermore, there was no statistically significant associations between sociodemographic characteristics, clinical staging, and post-operative complications. Mortality rate was 0%.

**Table 1:**
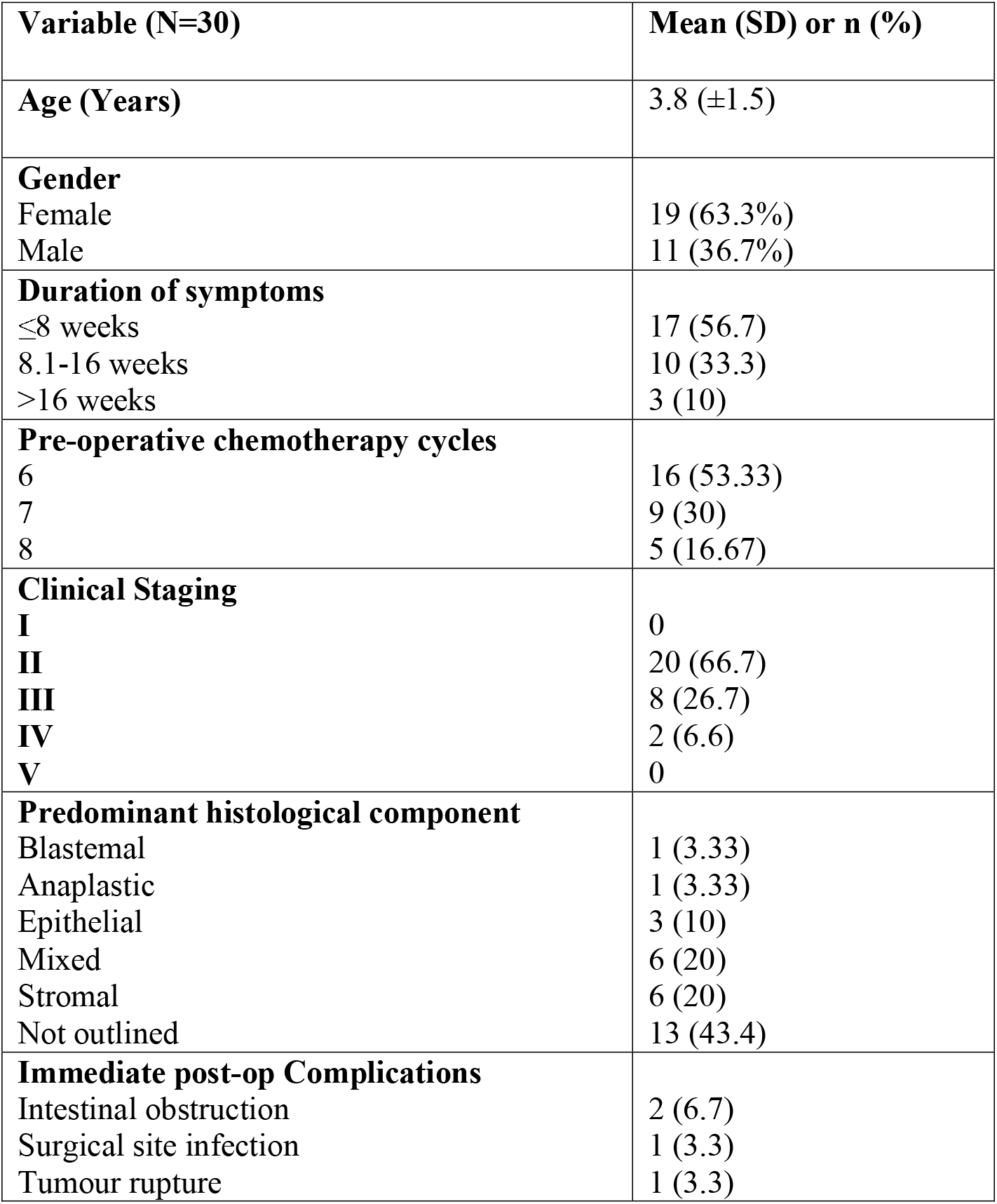
Demography and variables of the study subjects.

**Table 2:** Association between Duration of stay and other variables.

| Variable | Duration of stay |  | p-value |
| --- | --- | --- | --- |
| | N | $\rho$ | |
| Age | 30 | -0.090 | 0.634 |
| Chemotherapy sessions | 30 | 0.014 | 0.941 |
| Duration of symptoms | 30 | 0.396 | 0.044 |
| N-Population |  |  |  |
| ρ-Spearman Correlation | | | |

## Discussion

Wilms’ tumour represents one of the greatest success stories in modern medicine. The availability of the chemotherapy agents and antibiotic therapy have been major factors in the management of this tumour. However, surgical intervention is the mainstay of Wilms’ tumour management and remission. This study’s demographic findings compare well with those of previous studies done in Kenya ^6,7^, Malawi^8^ and China^9^.

However, the findings contrast those of a Nigerian study^10^ where the mean presentation age was 6 years. This could be attributed to the low awareness of the condition in some Sub-Saharan African countries, limited availability of chemotherapy agents and lack of proper Wilms’ tumour management mechanisms^11^. Poor health seeking behaviours among guardians of these patients and seeking alternative treatment options prior to getting medical advice could also have contributed to the late presentation.

All the children enrolled presented with abdominal distension and a palpable abdominal mass. Furthermore, nearly half of them had fever while less than one-tenth presented with macroscopic hematuria. This finding is comparable with that reported in Malawi^8^ where all patients presented with abdominal distension; and half of them had additional complaints including abdominal pain, haematuria, dyspnea, edema and or weight loss^8^. A large proportion (75%) of patients with abdominal mass was also reported in a Pakistani study^12^ alongside reports of fever, hematuria, weight loss among other symptoms. In a retrospective study conducted in Nigeria^11^, abdominal mass was the main presentation in all the children enrolled just as in this study.

The average duration of symptoms before admission was 8 weeks (IQR 4, 12) that was comparable to a Kenyan study^6^ where the majority of the patients presented within the first three months of symptoms onset. However, this finding contrasts two Nigerian studies^10,11^ where the duration of pre-clinical presentation was 4-14 months, and 4.7 months respectively. We attribute this late presentation to caretakers seeking other modes of treatment before finally seeking medical attention, ignorance, and poverty.

We further report that more than half (53.3%; n=16) of the participants received six cycles of neo-adjuvant chemotherapy before surgery, while 30% had seven cycles and 16.7% got 8 cycles. Neo-adjuvant chemotherapy has been demonstrated to consistently reduce the size of Wilms’ tumours. Studies conducted by the Society of Paediatric Oncology (SIOP) have reported a more than half mean reduction after a single cycle of chemotherapy and a further 50% reduction after the second cycle^13–15^. Furthermore, studies done in South Africa^16^ showed a mean reduction by 24% in tumour size following similar chemotherapy regimens. Neo-adjuvant chemotherapy plays an important role in complete surgical removal of a shrunken tumour, avoiding mutilation caused by surgical procedures and early treatment of micro-metastasis that were not visible at diagnosis^14,17,18^.

The early clinical outcomes of children with Wilms’ Tumor have significantly improved with combined therapeutic approaches integrating chemotherapy, surgery and radiotherapy^15^. The SIOP guidelines recommend that all Wilms’ Tumor patients should undergo pre-operative chemotherapy before surgery. This SIOP protocol has led to lower incidence of tumour rupture by reducing the vascularity of the tumour, risk of tumour spillage, a more favorable stage distribution and reduced treatment burden ^13, 14^. This study’s setting adopted the SIOP protocol to manage Wilms tumour’ in 2009. The use of SIOP protocols and the establishment of a multidisciplinary care team has led to improved management and better outcomes^6^. None of the children we enrolled died during the study period, however 13.3% of them had early complications.

Although early post-operative outcomes vary, local incidence of surgical outcomes following radical nephroureterectomy in Kenya has not been documented. Despite this, collaborative efforts among pediatric surgeons, pathologists, pediatric medical oncologists, and radio-oncologists have been regarded as a major factor in improving nephrouretectomy outcomes ^11^. The incidence of surgical complications reported in this study (13.3%) matched those of other trials, in the National Wilms’ Tumor Study-4 where complication rates of 12.6% were reported^19^.

Intestinal obstruction was the most common post-operative complications following radical nephroureterectomy among children with nephroblastoma. This study confirms previous reports that small bowel obstruction occurs between 5% to 12% among children with Wilms’ tumor^20^. In Germany, 8.82% of the children enrolled had to be reoperated because of small bowel obstruction^21^. In this study, the etiology of intestinal obstruction was small bowel intussusception, with the participants presenting with vomiting, abdominal pain and passage of red currant jelly stool three days after initiation of feeding.

Surgical site infection (SSI) is one of the post-operative complications that is associated with longer hospital stay, higher treatment costs and significant related morbidity and mortality. It includes superficial incisional, deep incisional, organ or space surgical site infections occurring in any area of the body other than skin, muscle and surrounding tissues that were involved in the surgery. In this study, 3.33% had surgical site infections that were treated through daily wound cleaning and dressing, and antibiotics administration. The proportion of wound infection reported in this study is similar to that in Canada where 2.6% of wound infections were reported^3^.

The goal of radical nephroureterectomy is complete resection of the tumour with tumour free margins and avoidance of intra-operative tumour rupture or spillage. This intra-operative spillage could increase risk of peritoneal seeding, upstaging of the tumour and a further recurrence. In this study, 3.3% rate of tumour rupture was reported intraoperatively due to surgical manipulation. In China, 4.5% of the patients had tumor rupture that could be attributed to fact that most patients presented in stage III^9^. However, much higher (21%) tumor spillage proportions were reported in South Africa^16^ that was attributed to the fact that the patients presented with large tumors.

The mean duration of stay in the surgical ward was 6.5 days which compared to a Pakistani study that reported 6.89 days^12^. Both studies were conducted in LMICs that may be experiencing similar health challenges. The duration of stay in the surgical ward was significantly associated with the duration of patient’s symptoms.

## Conclusions

The early surgical outcomes of children with Wilms’ tumour were favourable despite late presentation that increased the duration of stay in the surgical ward.

## Data Availability

Study data will be made publicly available.

## Acknowledgements

We would like to thank the Lecturers at Moi University School of Medicine and Consultant Surgeons at Moi Teaching and Referral Hospital who care and manage Wilms tumor patients. Furthermore, we would like to thank the hospital’s management for allowing us to conduct the study, the study participants and their parents for agreeing to take part in this study.

## Notes

### Competing Interest Statement

The authors have declared no competing interest.

### Funding Statement

This study did not receive any funding.

### Author Declarations

The study received prior ethical approval from the Moi University/Moi Teaching and Referral Hospital Institutional Research and Ethics Committee (IREC).

